# Investigating the relationship between serum vitamin D levels over time and the incidence of atrial fibrillation in The HUNT Study

**DOI:** 10.1101/2025.05.02.25326609

**Authors:** Lin Jiang, Yi-Qian Sun, Vegard Malmo, Xiao-Mei Mai

**Affiliations:** Department of Public Health and Nursing, Faculty of Medicine and Health Sciences, Norwegian University of Science and Technology, Trondheim, Norway; Clinic of Cardiology, St. Olav’s Hospital, Trondheim, Norway; Department of Clinical and Molecular Medicine, Faculty of Medicine and Health Sciences, Norwegian University of Science and Technology, Trondheim, Norway; Department of Pathology, Clinic of Laboratory Medicine, St. Olav’s Hospital, Trondheim University Hospital, Trondheim, Norway; TkMidt-Centre for Oral Health Services and Research, Mid-Norway, Trondheim, Norway; Department of Circulation and Medical Imaging, Norwegian University of Science and Technology, Trondheim, Norway

**Author notes:** Corresponding author Address for correspondence: Department of Public Health and Nursing, Faculty of Medicine and Health Sciences, Norwegian University of Science and Technology (NTNU), Postbox 8905, MTFS, N-7491 Trondheim, Norway.

**Keywords:** *Serum 25-hydroxyvitamin D*, *incidence of atrial fibrillation*, *The HUNT Study*, *repeated measurement*, *Mendelian randomization*

## Abstract

**Background and Aims:** Evidence of the association between serum vitamin D levels and atrial fibrillation (AF) is inconclusive. Thus, this study investigated the relationship between long-term average serum 25-hydroxyvitamin D (25(OH)D) levels and AF incidence in the Norwegian Trøndelag Health (HUNT) Study using a prospective cohort design and a Mendelian randomization (MR) approach.

**Methods:** A total of 3394 adults with two measurements of serum 25(OH)D at HUNT2 (1995-1997) and HUNT3 (2006-2008) and without AF at HUNT3 were followed up to 2021. Average serum 25(OH)D levels over ten years were categorized into <50 and ≥50 nmol/L. AF diagnoses were retrieved from hospital registers and validated by doctors. Cox regression was used to calculate hazard ratios (HR) and 95% confidence intervals (CI). Furthermore, a one-sample MR was conducted among 36,554 adults who participated in both HUNT2 and HUNT3 using the Wald ratio method.

**Results:** During a median 12-year follow-up, 304 AF cases were diagnosed. Serum 25(OH)D levels <50.0 nmol/L were associated with a 27% reduced incidence of AF (HR 0.73, 95% CI 0.57 to 0.93) compared with ≥50 nmol/L after adjustment for confounders. A genetically determined 10 nmol/L decrease in the serum 25(OH)D levels was associated with a 7% reduced incidence of AF (HR 0.93, 95% CI 0.86 to 1.00) in the one-sample MR. Sensitivity analyses supported this association.

**Conclusions:** Using both traditional observational and one-sample MR approaches, the study suggested a consistently positive association between long-term average serum 25(OH)D levels and incidence of AF in the Norwegian HUNT population.

## Introduction

Atrial fibrillation (AF) is the most commonly diagnosed arrhythmia and affects over 60 million people worldwide.^1^ It is an important risk factor for stroke, myocardial infarction and even sudden cardiac death.^2^ The prevalence of AF increases with age.^1^ Given the growing population of older adults globally, it is crucial to identify modifiable risk factors to prevent AF.^1^ Although the pathogenesis of AF is not fully understood, it is suggested that structural and electrical remodeling, regulation of the renin-angiotensin-aldosterone system (RAAS), inflammation and endothelial function are critical elements in the development of AF.^1,3^

Vitamin D is a micronutrient that is synthesized in the skin upon exposure to sunlight or obtained from dietary sources such as fatty fish and from supplements such as cod liver oil.^4^ Serum circulating levels of 25-hydroxyvitamin D (25(OH)D) are used to assess total vitamin D levels in the body since this measurement reflects the combined vitamin D obtained from sunlight exposure, dietary intake, and supplementation.^5^ Vitamin D insufficiency, typically assessed by a serum 25(OH)D concentration less than 50 nmol/L, affects over 50% of the world’s population, particularly during the winter season.^6,7^

Serum 25(OH)D has been suggested to play an important role in various physiological processes such as regulation of the aforementioned RAAS and inflammation.^8,9^ Therefore, it may be linked to the development of AF, but most previous prospective studies have not reported any associations.^10,11^ Since vitamin D levels fluctuate over time, a single measurement may not accurately represent a person’s long-term vitamin D status.^12^ As a result, a single measurement may lead to random measurement error and regression dilution bias in these studies.^13,14^ To better understand the relationship between serum 25(OH)D and the incidence of AF, more prospective studies that incorporate repeated measurements of serum 25(OH)D are needed.

During the past decade, Mendelian randomization (MR) method has been widely used to assist in drawing causal inference. The MR approach mimics Randomized Controlled Trial (RCT) in an observational setting by using genetic variants as instrumental variables for the risk factor of interest.^15^ Since genetic variants are randomly assigned at conception and remain stable over the lifetime, bias due to reverse causation may be avoided and the influence of residual confounding is reduced.^16^ It can offer supplementary evidence for causal relationships, while being less expensive and less time-consuming compared to RCTs.^15^ There is only one well-conducted two-sample MR study on this topic, which has not found any clear association.^17^ Compared with two-sample MR, one-sample MR benefits from a better control for confounders and reduced bias from population heterogeneity by utilizing individual-level data from a single population.^15^ To our knowledge, no one-sample MR has been conducted to investigate the potential association between serum 25(OH)D levels and AF incidence.

Hence, we first aimed to examine the relationship of average serum 25(OH)D levels over ten years with the incidence of AF in a prospective cohort study using the Norwegian Trøndelag Health (HUNT) population. Furthermore, we conducted a one-sample MR analysis to investigate the potential causal effect of serum 25(OH)D on AF incidence.

## Methods

### Study design and population

The HUNT Study is a large population-based health study that has been carried out in four phases, HUNT1 (1984-86), HUNT2 (1995-97), HUNT3 (2006-2008) and HUNT4 (2017-2019) in Trøndelag County, Norway.^18,19^ All adults aged 20 years or older were invited to complete general questionnaires on health and lifestyle status and undergo clinical examinations.^18,19^ In addition, information on the deaths and emigration of HUNT participants was regularly updated from the Norwegian National Registry.^18,19^

For our study, we included 37,069 adults who participated in both HUNT2 and HUNT3 (Figure 1). In the prospective cohort study, a random sample of the adult population with complete data on serum 25(OH)D levels in both HUNT2 and HUNT3 was included (n=3513). We followed them from HUNT3 (baseline) until the date of AF diagnosis, death, emigration or the end of follow-up (January 31, 2021), whichever occurred first. After excluding participants with a validated AF diagnosis before HUNT3 (n=119), 3394 participants were left in the prospective cohort study.

**Figure 1.**
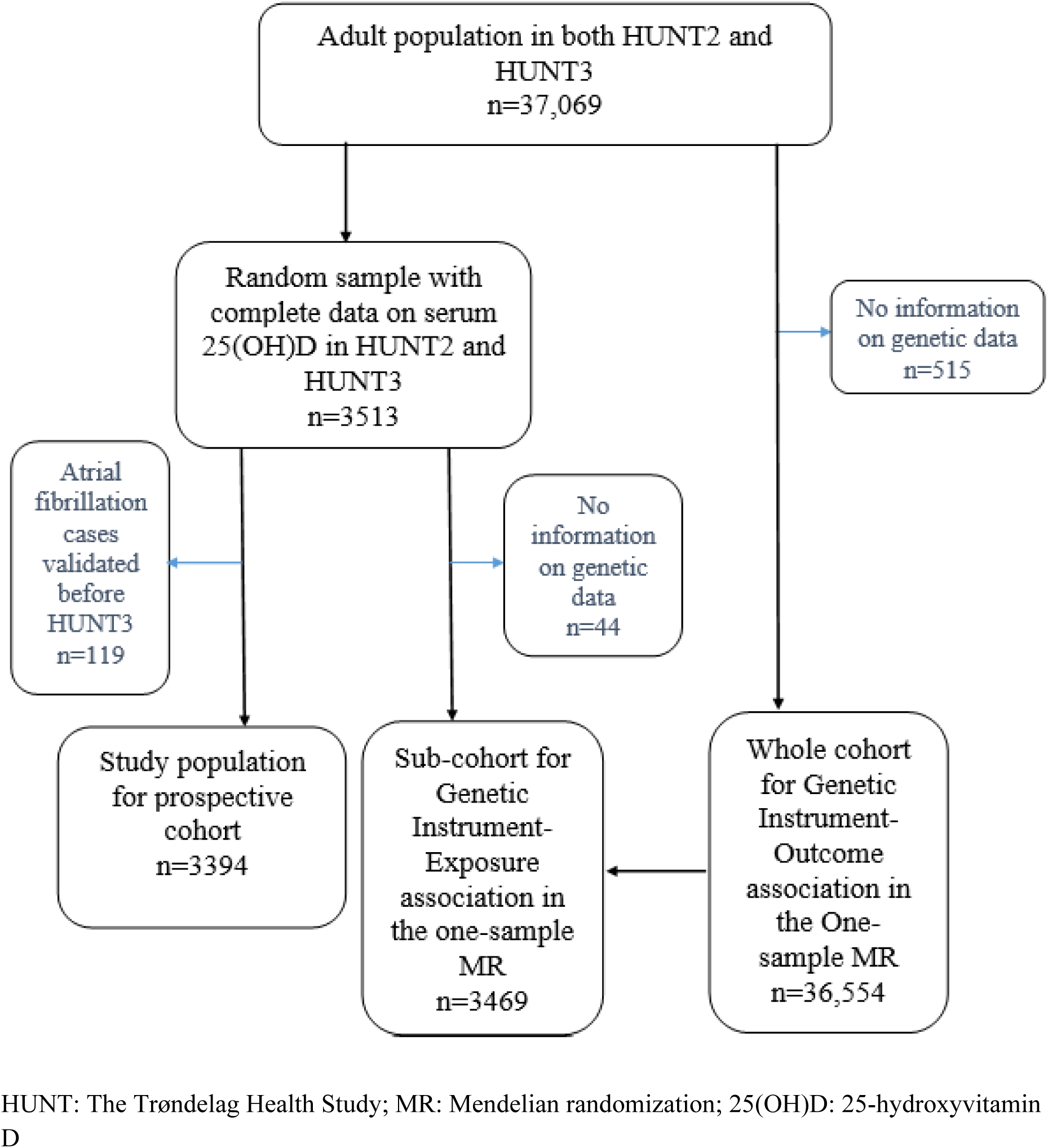
Flow chart of study populations

Of the 3513 individuals with complete data on serum 25(OH)D levels in both HUNT2 and HUNT3, 3469 had information on genetic data and were included as a sub-cohort in the one-sample MR to estimate the genetic instrument-exposure association. The cohort used for estimating the genetic instrument-outcome association (n=36,554) consisted of participants with complete genetic data derived from the 37,069 adults who participated in both HUNT2 and HUNT3 (Figure 1).

### Measurements and standardization of serum 25(OH)D levels

Serum 25(OH)D levels in HUNT2 and HUNT3 were measured at the HUNT Biobank using the LIAISON 25-OH Vitamin D TOTAL (DiaSorin, Saluggia, Italy), a fully automated, antibody based, chemiluminescence assay.^20^ The detection range of the assay for total serum 25(OH)D is 10-375 nmol/L. Measurements of serum 25(OH)D were standardized to account for seasonal variations.^21^ The standardized values reflect serum 25(OH)D concentrations measured in October for each individual,^21^ effectively addressing seasonal fluctuations in the levels caused by the high latitude of Norway. The long-term average serum 25(OH)D levels were calculated as the mean of the two measurements in HUNT2 and HUNT3. For simplicity, the average season-standardized levels over 10 years are termed as the serum 25(OH)D levels thereafter in this study. We categorized serum 25(OH)D levels into four initial groups: <30.0, 30.0-49.9, 50.0-74.9 and ≥75.0 nmol/L based on the definition by the Institute of Medicine^7^ and then combined the first two groups as <50.0 nmol/L to represent insufficient levels. The remaining two groups were combined as ≥50.0 nmol/L to represent sufficient levels.

### Vitamin D SNPs and the genetic risk score as instrumental variables

Extracted DNA from blood samples collected at HUNT2 or HUNT3 was stored in the HUNT Biobank. Genome-wide genotyping and imputation were carried out with sample and variant quality control by using Illumina Humina HumanCoreExome arrays.^22^ We utilized 21 single nucleotide polymorphisms (SNPs) from four gene regions (GC, DHCR7, CYP2R1 and CYP24A1) as candidate instrument variables for serum 25(OH)D, as reported by Sofianopoulou et al..^21^ These SNPs were selected based on their well-established biological roles in vitamin D transport, synthesis and metabolism,^21^ and therefore they had strong associations with serum 25(OH)D levels (P value <5×10^-8^).

Information on two SNPs (rs139148694 and rs35870583) was missing from the HUNT data because they did not pass imputation quality control (R^2^ of linkage disequilibrium >0.8). Consequently, 19 SNPs were used to construct an externally weighted genetic risk score (GRS) for our analyses. Using a GRS instead of individual genetic variants can ensure that a large proportion of serum 25(OH)D can be accounted for and therefore reduce weak instrument bias and increase statistical power.^23^ The externally weighted GRS was calculated as the sum of the number of effect alleles carried for each SNP weighted by the reported beta coefficient (β) for serum 25(OH)D derived from the study by Sofianopoulou et al..^21^ The characteristics of the 19 individual SNPs are shown in Supplementary Table 1.

### Other baseline variables

In HUNT3, body weight, height and blood pressure were measured by health professionals at the clinical examination. Height was measured to the nearest centimeter and weight to the nearest 0.5 kg. Body mass index (BMI) was calculated as weight in kilograms divided by height squared in meters (kg/m^2^). Other covariates included: sex (women and men), smoking status with detailed pack-years (pyrs) information [(never, former (<10, 10-20 and >20 pyrs) and current (<10, 10-20 and >20 pyrs)], alcohol consumption (never, 1-4 and ≥5 times/month), physical activity (inactive, low, moderate and active), occupation status (Class I to VII), hypertension (yes and no), coronary heart disease (yes and no), diabetes (yes and no), serum creatinine (µmol/L) and serum cholesterol (<5.2 nmol/L as desirable, 5.2-6.2 nmol/L as borderline and >6.2 nmol/L as high). Hypertension was defined as systolic blood pressure ≥140 mmHg or diastolic blood pressure ≥90 mmHg or self-reported use of antihypertensive medication.^24–26^ Coronary heart disease (CHD) was defined based on a “yes” answer to questions about angina pectoris or heart attack history. Diabetes was defined based on a “yes” answer to question about diabetes history and/or the non-fasting blood glucose levels above 11 mmol/L. Missing information on each of the mentioned variables was included in the analyses as an “unknown” category. We used the same categorization of variables as in previous HUNT publications.^12,27^

### Verified AF diagnosis

AF diagnoses were retrieved from discharge registers at two hospitals in the North Trøndelag County using the unique 11-digit personal identification numbers. The diagnosis was based on the International Classification of Diseases 10th revision (ICD-10) code I48 for atrial fibrillation/atrial flutter.^28^ The verified AF diagnosis was AF or atrial flutter confirmed by a physician after reviewing the electrocardiograms (ECGs) according to standard criteria.^1^ To confirm AF cases among participants with physician-diagnosed AF but no ECG information, two physicians independently assessed the available data, and the “probable AF” cases agreed by both physicians were treated as verified AF.^28^ Persons who only had an episode of AF within the first 7 days after cardiac surgery, during the acute phase of a myocardial infarction or during episodes of severe hemodynamic instability (e.g. sepsis or major non-cardiac surgery) were not regarded as having incident AF.^28^

### Statistical analysis

The baseline characteristics of the participants included in the prospective cohort study (n=3394) and the one-sample MR study (n=36,554) are presented. The P value for the test of linearity was less than 0.05, which indicated that the association between the serum 25(OH)D levels and the incidence of AF might have deviated from a linear association. Thus, we only treated serum 25(OH)D levels as a categorical variable in the analysis. Cox proportional hazard models were used to examine the potential associations. We assessed the proportional hazards assumption by Schoenfeld residuals for exposure and all covariates. The results suggested that the proportional hazard assumption held for both exposure and covariates. Crude and adjusted hazard ratios (HRs) with 95% confidence intervals (CIs) were calculated with age as the underlying time scale. Potential confounders were selected based on previous knowledge ^29–31^ and directed acyclic graph (DAG). In the main model, we adjusted for sex, BMI, smoking status, alcohol consumption, physical activity, occupation, hypertension and CHD. Diabetes, serum creatinine and serum cholesterol status were adjusted in an additional model since they may be mediators of the association.^30,31^

To test the robustness of the findings in the prospective cohort analysis, we performed a series of additional analyses: 1) Classified the serum 25(OH)D levels into four categories (<30.0, 30.0-49.9, 50.0-74.9 and ≥75.0 mmol/L)^7^ to gain a more detailed understanding of its relationship with AF. 2) Excluded the first three-year follow-up and participants with CHD at baseline to address reverse causality due to existing but undiagnosed AF. 3) Conducted multivariable chained imputation with fully conditional specification (m=10 imputed datasets) for missing information of all covariates to address residual confounding. 4) Performed a positive outcome control analysis using diabetes as an outcome^32^ in the prospective cohort population to investigate potential selection bias.^33^ Meta-analyses of observational studies and RCTs have consistently suggested an inverse association between serum 25(OH)D and diabetes risk.^34^ If we observe the same direction of association, selection bias may not be a major methodological issue in our study population. 5) Performed a competing risk analysis based on the Fine-Gray model to address possible competing risk due to death.^35^

Furthermore, we conducted a one-sample MR analysis to assess the potential causal association between genetically predicted serum 25(OH)D levels (per 10 nmol/L decrease) and the incidence of AF. Baseline characteristics were compared between the population for the GRS-outcome association (n=36,554) and its sub-cohort for the GRS-exposure association (n=3469).^15,36^ A Wald ratio method was applied to compute the MR estimates.^15,37^ We computed a MR-driven hazard ratio by applying the natural exponential function of the ratio of coefficients. All regression models were adjusted for age, sex, batch and 20 PCs.^21^

In the sensitivity analysis of the one-sample MR approach, we first tested the relevance assumption using the F statistic and R^2^ value for the association between the GRS and serum 25(OH)D levels in the sub-cohort of 3469 individuals. The GRS is regarded as an adequate instrument variable if the F-statistic >10.^38^ Next, we tested the associations between the GRS and the available confounders collected in HUNT3 at baseline among the 36,554 individuals using linear or logistic regressions. The GRS should not be associated with any potential confounders according to the independence assumption. We further assessed the exclusion assumption using SNP-based two-sample methods such as the inverse-variance weighted method (IVW), MR-Egger, simple median, weighted median ^39,40^ and MR-PRESSO ^41^ methods.

All the statistical analyses were performed with STATA/SE 16.1 (College Station, TX, USA) or R (4.3.2). The package “TwoSampleMR” was used for the SNP-based two-sample methods in R.

## Results

Table 1 describes that the distributions of all baseline characteristics were similar in the prospective cohort (n=3394), the sub-cohort for genetic instrument-exposure association (n=3469) in the one-sample MR and the entire cohort for the genetic instrument-outcome association (n=36,554).

**Table 1.**
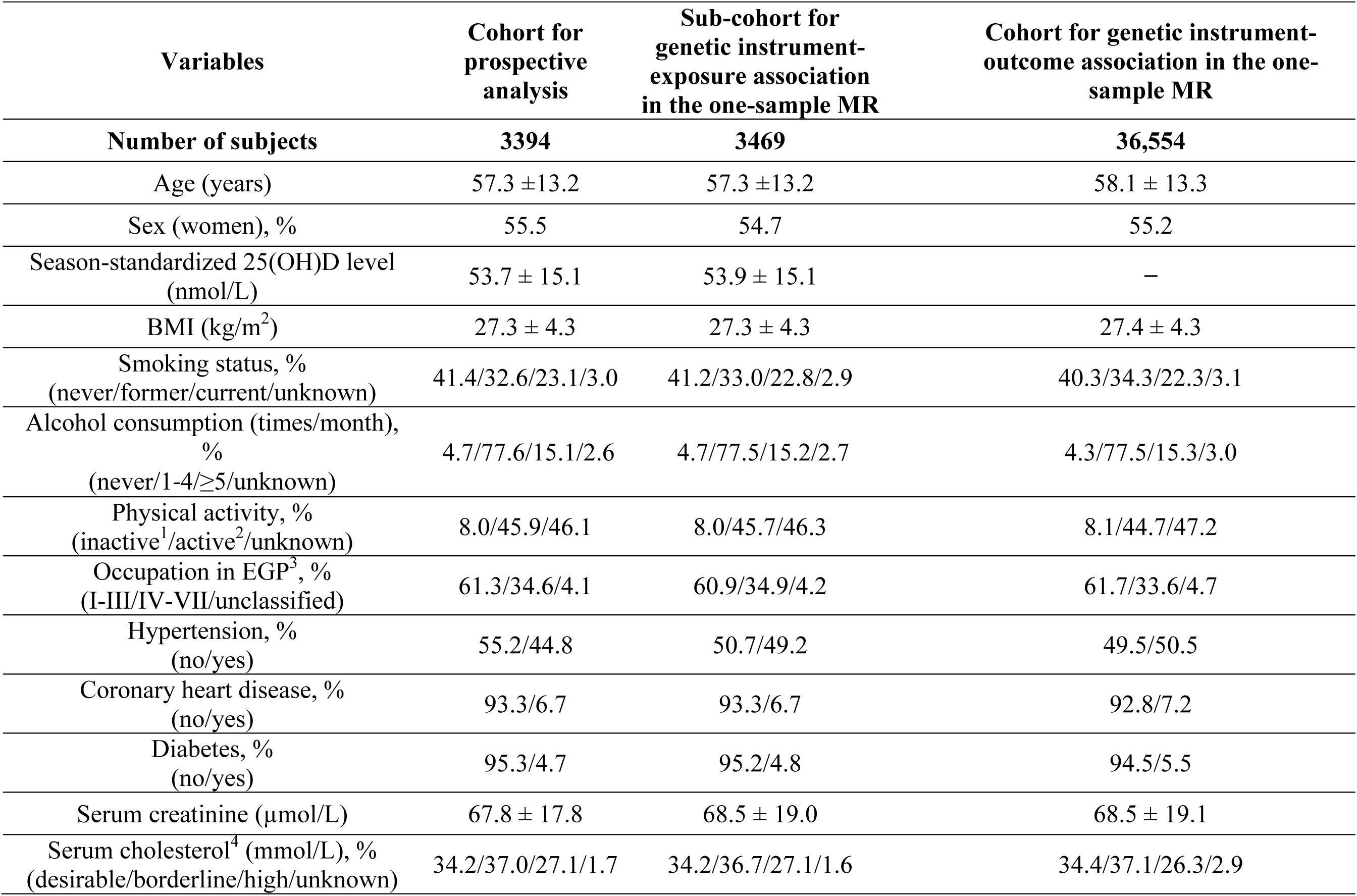
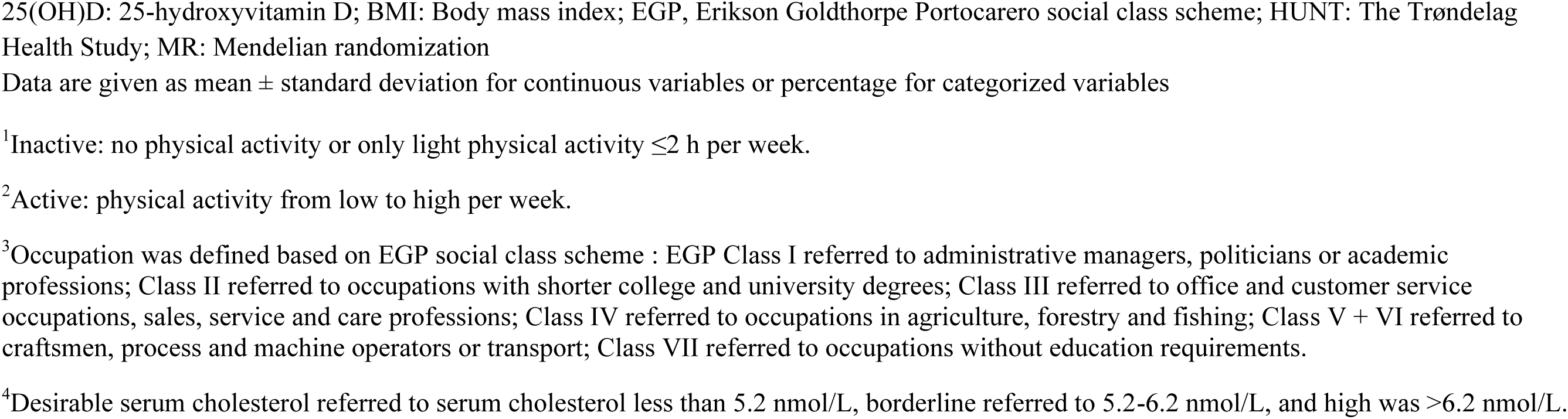
Baseline (HUNT3) characteristics of participants included in the prospective cohort and the one-sample MR analyses.

In the prospective cohort study, we first compared the baseline characteristics of adults across their serum 25(OH)D categories (Supplementary Table 2). Compared with those with serum 25(OH)D levels ≥50.0 nmol/L, participants with serum 25(OH)D levels <50.0 nmol/L seemed to have higher percentage of obesity, current smoking (>20 pack-years), hypertension, diabetes, and elevated cholesterol levels, as well as lower levels of moderate to high physical activity and lower percentage of occupational classes related to high social class. Among the 3394 individuals, 304 developed AF during a median follow-up of 12.1 years. Serum 25(OH)D levels <50.0 nmol/L were associated with a 27% reduced incidence of AF after adjustment for confounders in the main model (HR 0.73, 95% CI 0.57 to 0.93), compared with serum 25(OH)D ≥50.0 nmol/L (Table 2). Additional adjustments for diabetes, serum creatinine and cholesterol levels did not change the results.

**Table 2.** Prospective association of average serum 25(OH)D levels between HUNT2 and HUNT3 with the incidence of atrial fibrillation, the HUNT Study, 2006-2008 to 2021 (n=3394)

|  | Cases | IR (per 1000 person-years) | Crude<sup>1</sup> |  | Main model<sup>2</sup> |  | Additional model<sup>3</sup> |  |
| --- | --- | --- | --- | --- | --- | --- | --- | --- |
|  |  |  | HR | 95% CI | HR | 95% CI | HR | 95% CI |
| **Average serum 25(OH)D between HUNT2 and HUNT3** | 304 |  |  |  |  |  |  |  |
| <50.0 nmol/L | 114 | 6.25 | 0.78 | 0.62-0.98 | 0.73 | 0.57-0.93 | 0.73 | 0.57-0.93 |
| ≥50.0 nmol/L | 190 | 8.31 | 1.00 | Reference | 1.00 | Reference | 1.00 | Reference |

25(OH)D: 25-hydroxyvitamin D; CI: confidence interval; HR: hazard ratio; HUNT: The Trøndelag Health Study; IR: Incidence rate
<sup>1</sup>Age was used as the time scale in the crude model.
<sup>2</sup>In the main model, we adjusted for sex, body mass index (as continuous), smoking [(never, former (<10, 10-20, and >20 pack-years (pyrs)), current (<10, 10-20, and >20 pyrs)], alcohol consumption, physical activity, occupation, hypertension and coronary heart disease. Age was used as the time scale.
<sup>3</sup>Additional adjustment for diabetes, serum creatinine, and cholesterol status in addition to the covariates in the main model.

The results of additional analyses provided supportive evidence for our primary findings: 1) Findings derived from four categories of serum 25(OH)D levels seemed to align with our primary results but with broader 95% CIs. However, the results did not demonstrate a linear dose-response association (Supplementary Table 3). 2) Excluding the first three years follow-up and participants with CHD yielded similar results (Supplementary Table 4). 3) Multiple imputation for all covariates with missing showed comparable association estimates (Supplementary Table 5). 4) In the analysis using diabetes as a positive outcome control, serum 25(OH)D levels measured at HUNT2 were inversely associated with the risk of diabetes at HUNT3 (Supplementary Table 6), which is in line with the results of previous studies.^34^ This finding indicated that selection bias was unlikely to be the reason for our observed positive association between 25(OH)D and AF incidence. 5) Competing risk due to death did not influence our primary results (Supplementary Table 7).

In the one-sample MR, a genetically determined 10 nmol/L decrease in serum 25(OH)D was associated with a 7% reduction in AF incidence (HR 0.93, 95% CI 0.86 to 1.00) using the Wald ratio method (Table 3). Sensitivity analyses using SNPs-based two-sample methods such as IVW, MR-Egger, simple median and weighted median methods generally supported our primary causal findings (Table 3).

**Table 3.** Associations of genetically predicted serum 25(OH)D levels (per 10 nmol/L decrease) with the incidence of atrial fibrillation in the one-sample MR analysis (n=36,554)

|  | **Methods** | **HR** | **95% CI** | **P value** |
| --- | --- | --- | --- | --- |
| **GRS-based one-sample MR analysis** | Wald ratio | 0.93 | 0.86 to 1.00 | 0.04 |
| **SNPs-based two-sample MR methods** | IVW | 0.92 | 0.85 to 0.98 | 0.02 |
|  | MR-Egger | 0.94 | 0.83 to 1.06 | 0.31 |
|  | MR-Egger Intercept | Effect size |  |  |
|  |  | 0.01 | -0.02 to 0.04 | 0.63 |
|  | Simple median | 0.89 | 0.77 to 1.04 | 0.14 |
|  | Weighted median | 0.94 | 0.87 to 1.02 | 0.16 |

25(OH)D: 25-hydroxyvitamin D; CI: Confidence interval; GRS: Genetic risk score; HR: Hazard ratio; HUNT: The Trøndelag Health Study; IVW: Inverse-variance weighted method; MR: Mendelian randomization; MR-PRESSO: Mendelian Randomization Pleiotropy RESidual Sum and Outlier; SNPs: Single nucleotide polymorphisms

In addition, the GRS explained 8.6% of the variability in serum 25(OH)D levels among the sub-cohort of 3469 individuals, with an F-statistic of 324. The GRS did not appear to be associated with any of the measured confounders among the 36,554 individuals (Supplementary Table 8). Cochran’s Q tests suggested no heterogeneity (P for Q >0.05, results not shown). The intercepts from the MR-Egger method did not deviate from zero and the P value for the intercept was 0.63 (Table 3). Thus, the MR-Egger method did not show evidence of a directional pleiotropic effect under the Instrument Strength Independent of Direct Effect assumption (InSIDE).^15^ No outliers were detected based on the results from the MR-PRESSO method.

## Discussion

We observed a positive association between serum 25(OH)D levels, representing the long-term average value over ten years, and the incidence of AF after a median follow-up of 12 years in a prospective cohort study of the Norwegian adult population. Our one-sample MR provided further evidence supporting this positive association.

Majority of the previous prospective studies did not suggest a clear association between serum 25(OH)D levels and the incidence of AF among the general population.^10,11^ Most of the studies included participants older than 65 years.^10^ In addition, only one single measurement of serum 25(OH)D was used, and AF cases were not verified in most of the previous studies.^10,11^ The measurement errors of exposure and outcome might give biased results. In our prospective study, we addressed these issues by using repeated measurements of serum 25(OH)D and verified AF diagnosis. This might be an explanation for the difference in findings between our and the previous prospective studies.

We further found a positive association between a genetically determined 10 nmol/L decrease in the serum 25(OH)D levels and a reduced incidence of AF in our one-sample MR. However, we need to interpret this finding cautiously since no association was found in a previous two-sample MR including the largest number of AF cases (cases/controls=60,620/970,216).^17^ Nevertheless, the two-sample MR used summary data for AF from six different cohorts, which might increase bias due to heterogeneity. Moreover, unlike the use of individual genetic variants as instruments in the two-sample MR, we used an externally weighted GRS that enhanced our instrument strength. In addition, we measured serum 25(OH)D twice and the long-term average value might offer a more reliable estimate for the exposure and reduce the likelihood of Type II errors.

Furthermore, mixed evidence on causality has been reported based on available RCTs with shorter follow-up durations.^42,43^ The Vitamin D and Omega-3 Trial (VITAL) Rhythm Study suggested no effect of vitamin D supplementation on the prevention of AF at 2000 IU/day over 5 years among 25,119 healthy adults.^42^ The included participants had relatively high baseline serum 25(OH)D levels of approximately 75 nmol/L. This might diminish the response to supplementation and subsequently lead to null finding. In another Finnish RCT over 5 years among 2495 individuals, the authors found a potential benefit in preventing AF using a vitamin D supplementation dose similar to that used in the VIVAL Rhythm Study.^43^ However, the beneficial effect was obtained from a post hoc analysis, and a possibility of a chance finding cannot be ruled out in such a small RCT study.

Our study has several strengths. First, this is the first study using both the prospective cohort study and a one-sample MR approach to investigate the potential association between serum 25(OH)D levels and AF incidence. Second, our study benefits from a relatively long follow-up duration over 12 years. Third, selection bias is likely minimal, as the HUNT study had a relatively high participation rate compared with other studies.^18^ Our study cohorts for the prospective analysis and the GRS-exposure association are fairly representative of the entire population used for the GRS-outcome association. In addition, our results using diabetes as a positive outcome control further confirmed that selection bias was not a major issue in this study. Fourth, measurement errors of exposure were better controlled by measuring serum 25(OH)D twice and using the average values to reflect its levels over time. The verified AF cases from the discharge registers demonstrated greater validity than those from the ICD codes alone.^28^ Fifth, we had information on a panel of potential confounders at baseline, which could minimize confounding. Finally, using the one-sample MR approach with individual-level data, we had the opportunity to investigate the three MR assumptions: 1) The instrument, composed of 19 SNPs with well-established biological roles in vitamin D transport, synthesis and metabolism, explained 8.6% of the variability in serum 25(OH)D levels, suggesting its suitability of being an appropriate instrumental variable. 2) We were able to investigate the associations between the GRS and a panel of potential confounders including lifestyle factors and clinical data. 3) There was no substantial violation of the exclusion assumption since we did not observe horizontal pleiotropy based on the results from the MR-Egger and MR-PRESSO methods, although pleiotropy could manifest in subtle ways.

Our study had several limitations. First, in our prospective cohort analysis, we cannot completely rule out the possibilities of residual confounding. The latest RCTs suggest that omega-3 supplements may be associated with an increased incidence of AF.^44–46^ Since many Norwegians might obtain vitamin D supplementation through cod liver oil, which contains Omega-3, Omega-3 likely acted as a positive confounder in our observational analysis. However, the observed positive association between serum 25(OH)D and AF incidence remained unchanged after further adjustment for cod liver oil or Omega-3 supplements (data not presented). Second, compared with the previous two-sample MR study,^17^ the sample size in our one-sample MR was smaller.^17^ Nevertheless, our genetic instrument strength appeared to be stronger than the instrument strength in the two-sample MR study (F statistics: 324 in our study vs a range of 13-254 in previous studies). The discrepancy in findings underscores the need for more one-sample MR studies with individual-level data and larger sample sizes in the future. Third, we were unable to investigate the potential causal non-linear association between serum 25(OH)D and AF incidence. This is because 25(OH)D levels are not measured across the entire HUNT population, and widely accepted methods for non-linear MR analyses are currently not available.^47^ Last, although the HUNT population is a homogeneous population with over 97% Caucasians, the homogeneity may restrict the generalizability of the findings to other ethnic groups.

## Conclusion

We observed a positive association between long-term average serum 25(OH)D levels and the incidence of AF. The results from the one-sample MR approach supported this association. More one-sample MR studies with larger sample sizes are needed to clarify this association. Until then, clinical and public health recommendations regarding vitamin D supplementation for the prevention of AF risk should be made cautiously.

## Data Availability

Data from the HUNT Study that is used in research projects will, when reasonably requested by others, be made available on request to the HUNT Data Access Committee.

https://www.ntnu.edu/hunt/data

## Acknowledgments

The HUNT Study is collaboration between HUNT Research Centre (Faculty of Medicine and Health Sciences, Norwegian University of Science and Technology - NTNU), Nord-Trøndelag County Council, Central Norway Regional Health Authority and the Norwegian Institute of Public Health. The genotyping in HUNT was financed by the National Institutes of Health; University of Michigan; the Research Council of Norway; the Liaison Committee for Education, Research and Innovation in Central Norway; and the Joint Research Committee between St Olavs hospital and the Faculty of Medicine and Health Sciences, NTNU.

## Authors’ contributions

VM was responsible for data collection. LJ conducted statistical analyses, interpreted results and wrote the initial draft of the manuscript. YQS and XMM contributed to the study design and statistical analyses. VM, YQS and XMM participated in the data interpretation, contributed to the manuscript writing with important intellectual content. All authors approved the final version of the manuscript.

